# ADVANCED AUTOLOGOUS LOWER DERMAL SLING TECHNIQUE FOR IMMEDIATE BREAST RECONSTRUCTION SURGERY IN SMALL AND NON-PTOTIC BREASTS

**DOI:** 10.1101/2021.05.09.21255654

**Authors:** Chaitanyanand B. Koppiker, Aijaz Ul Noor, Santosh Dixit, Laleh Busheri, Gautam Sharan, Upendra Dhar, Hari Kiran Allampati, Smeeta Nare, Nutan Gangurde

## Abstract

**Background:** Breast reconstruction with an autologous lower dermal sling (ALDS) is an established one-stage procedure in patients with moderate to large ptotic breasts. However, this technique is difficult to perform in small and non/minimally ptotic breasts. We describe our experiences from a single institution about a novel Advanced Autologous Lower Dermal Sling (A-ALDS) technique for reconstruction in small breasts.

**Methods:** We performed one stage nipple/skin sparing mastectomies in 61 patients with immediate reconstruction either by conventional immediate breast reconstruction surgery or A-ALDS technique.

**Results:** Mean age of study patients was 46.9 years. We observed significantly better cosmetic score and lower immediate complication rate *vis-a-vis* skin necrosis, implant loss with the A-ALDS technique (i.e., nil versus 3 in Conventional Immediate Breast Reconstruction Surgery - IBRS). 40 patients completed 12 months follow-up. The PROMs-Patient Reported Outcomes Measures (Breast-Q) revealed good to excellent scores for satisfaction with breast, cosmetic outcome and psychosocial well-being in patients operated with both these techniques. However, sexual well-being was significantly better in the A-ALDS group.

**Conclusion:** The A-ALDS is a novel, cost-effective and safe technique for immediate one stage implant-based reconstruction for small breasts. It provides a dermal barrier flap and hence, ensures less complications, excellent cosmetic results and patient satisfaction.

## INTRODUCTION

The well-established autologous lower dermal sling (ALDS) is an ideal technique for implant-based immediate breast reconstruction surgery (IBRS) in breast cancer (BC) patients with moderate-to-large sized breasts with significant ptosis (Grade 2+) in which the distance from the nipple to the infra-mammary fold (IMF) may be significantly greater than 5-7 cm. The ALDS provides robust stability to the implant ensuring excellent contouring, projection and fullness of the lower pole of the reconstructed breast with correction of ptosis. This approach reduces the risk of a high riding implant, provides excellent cosmetic results and ensures good symmetry. Therefore, ALDS is now considered as a safe and cost-effective single stage reconstructive technique especially in moderate-to-large ptotic breasts [1–3].

However, implant-based breast reconstruction for small and non-ptotic breasts is a surgical challenge due to non-availability of excess lower pole length that aids in the creation of an appropriately sized dermal sling. Conventionally, breast reconstruction in such scenarios is performed by placing the implant either in a complete or partial sub-muscular pocket. These techniques have been shown to result in sub-optimal cosmetic outcomes that were attributed to poor expansion and tightness of the inferior pole [4]. As a result, it is now a common practice employed by several oncoplastic surgeons to use acellular dermal matrices (ADMs) that function as a sling to cover the implant inferiorly and provide robust support to the inferior pole [4].

In developing countries such as in India, ADMs are not available and their high cost can prohibit its use in breast reconstruction. Hence, in such low-resource settings, we have initially approached reconstruction in minimally ptotic or non-ptotic small breasts by placing the implant in a sub-muscular pocket that is formed by pectoralis major muscle above, the fascia or superficial fibers of the serratus anterior muscle laterally and the fascia over rectus abdominis muscle inferiorly. Later, we developed a technique of Advanced-Autologous Lower Dermal Sling (A-ALDS) to provide a double layer of vascularized tissue cover over the implant inferiorly. It has been previously reported in the ALDS procedure, that dermal sling provides an advantage over ADMs as it acts as a vascularized flap eliminating the risk of implant exposure even if superficial skin necrosis does occur [1–2]. Therefore, the ALDS technique has been utilized routinely to improve breast reconstruction outcomes. Similarly, our modified A-ALDS technique was able to maintain the natural breast shape, projection and symmetrization of the reconstruction with respect to the opposite breast thereby, obviating the need for contralateral surgery.

In this study, we report the application of this novel A-ALDS technique for reconstruction in patients with small breasts. Furthermore, we present the post-surgical outcomes in our study patients who have undergone either A-ALDS technique or conventional IBRS.

We present the following article/case in accordance with the STROBE reporting checklist.

## MATERIAL AND METHODS

### Study Design

This is a longitudinal cohort study involving a retrospective analysis of prospective data from a single institution. This study was approved by an independent ethics committee associated with the institution. Patients were considered eligible for analysis if they underwent unilateral and bilateral mastectomies with implant-based IBRS (Immediate Breast Reconstruction Surgery) and fulfilled the criteria laid down for a small breast. Small breast for the purpose of this study was defined as the one with a cup size of B or smaller and / or mastectomy weight of less than 350gm with either no or normal ptosis.

Written informed consent was obtained from all patients for collection of study-relevant medical data associated with disease management and routine follow-up visits. The study recruitment period was defined as the day informed consent was obtained from the patient till one year of follow up. Study sample size represented all eligible cases during the study period from year 2016 until 2018. Data collection included demography, medical history, clinicopathological characteristics, surgical notes, post-surgery evaluations and follow-up details for patient reported outcome measures (PROMs) and aesthetic scores. To minimise bias in data collection, Standard Operating Procedures (SOPs) were implemented by well-trained researchers.

During the study period, a total of 61 BC patients with small breasts underwent implant-based IBRS at our breast unit. 3 patients underwent bilateral mastectomies. Only those patients with small breasts who were recommended for mastectomy and opted for IBRS were included in the study. The selection criteria for A-ALDS (Advanced-Autologous Lower Dermal Sling) or conventional IBRS as an appropriate surgical technique were as follows: (a) Conventional IBRS was performed when the tumor was located in the upper outer quadrant and lower quadrant close to the skin, warranting skin removal with the tumor. (b) The A-ALDS procedure for IBRS was applied to all other patients including those that had the presence of tumor in the lower pole but at an optimal distance from skin. In such situations, the skin could be preserved to create the desired dermal sling. The surgical margins especially – the anterior margins were evaluated by frozen sections in selected cases and re-ascertained on the paraffin sections in all cases.

Out of these 61 patients, 40 completed one-year post-surgery follow-up (22 with conventional IBRS and 18 with A-ALDS) and were analyzed for surgical outcomes and PROMs. These patients underwent chemotherapy and/or radiation therapy treatment according to NCCN guidelines under the supervision of a multidisciplinary clinical team.

### Conventional IBRS Technique

Conventional IBRS (i.e.one stage sub muscular implant reconstruction) involves placement of the breast implant in a sub muscular pocket. The mastectomy is performed preserving the skin along with nipple areola complex in which the lower extent of mastectomy is the inframammary fold (IMF). The conventional IBRS technique involves splitting pectoralis major muscle in the middle along its fibers. This is in contrast to the usual practice of most plastic surgeons that involves lifting the pectoralis major starting from its lateral edge and then dissecting beneath it to create a sub-muscular pocket. Laterally, the dissection is carried under the fascia/superficial fibers of serratus anterior, hence providing a continuous pocket laterally. The dissection then continues under the fascia of lower thoracic and upper abdominal wall approximately 2 cm below IMF. This procedure ensures the provision of an appropriate cover to the implant to prevent high riding and imparts fullness to the lower quadrant of the reconstructed breast. In this way, the implant is partially covered by muscle and partially by fascia. We have adopted a single stage procedure by inserting a dual lumen expendable implant so as to create lower pole fullness.

### Advanced-Autologous Lower Dermal Sling (A-ALDS) Technique

The A-ALDS procedure is described for implant-based reconstruction in patients with small non-ptotic or minimally ptotic breasts where the distance of the lower segment of the breast is inadequate to perform the regular ALDS technique.

The A-ALDS technique begins by first marking out the usual pre-operative landmarks on the breast with appropriate measurements. The IMF is marked and the IMF-to-nipple distance is ascertained (typically small; 5 to 7 cm). The desired dermal sling (breadth of 3 to 4 cm) is marked out in a semi-circular fashion above the IMF on the lower pole of the breast. This area is de-epithelialized to constitute the lower dermal sling. The mastectomy proceeds from above the lower dermal sling and the flaps are raised in the appropriate subcutaneous plane maintaining the sub-dermal blood supply of the flaps. At the areola, the plane becomes more superficial and then dips into the nipple to core out the ducts as recommended [5]. The lower dermal sling is dissected off the lower breast tissue in the correct plane up to the IMF. Then, mastectomy is performed and the nipple is cored out. During this procedure, the lower dermal sling is then advanced by mobilizing the skin and subcutaneous tissue above the fascia of the lower thoracic and upper abdomen by 3 to 4 cm thereby, advancing the dermal sling to cover the implant. We recommend that during this mobilization the perforators of medial and lateral thoracic region should be maintained scrupulously as they contribute significantly to the vascularity of A-ALDS flap. The advanced skin mobilized (from lower thoracic and upper abdomen) equals the breadth of the dermal sling (i.e. 3 to 4 cm), maintains the required ideal length of the lower segment and helps to symmetrize the reconstructed breast to the contralateral side. Finally, the IMF is recreated by anchoring the mobilized skin (from the lower thoracic and upper abdominal wall) with 3 sub-cutaneous sutures to the chest wall at the level of the original IMF.

After this step, a breast pocket is created by lifting up the pectoralis major muscle from the chest wall and cutting its attachments inferiorly. The pocket is continued under the fascia and superficial fibers of the serratus anterior muscles without detaching the pectoralis major muscle at its lateral attachment providing a continuous uninterrupted pocket laterally. Medially, the dissection is carried under the pectoralis major muscle up-to the medial perforators without damaging them. An appropriate size and type of dual-lumen anatomical implant is placed in the resultant submusculardermal pocket. To avoid excess tension inside the submuscular pocket, the volume of the pocket is kept proportionate to the skin envelope and to implant dimension.

In the final surgical step, the inferior border of pectoralis major muscle and superficial fibers of serratus anterior or its fascia muscle is sutured to the de-epithelialized A-ALDS generated earlier. The suction drains are placed under skin flaps of the patient and the skin flap is sutured down at the infra-mammary crease (Figure 1A a-e) (Online Resource 1-3).

**Figure 1:**
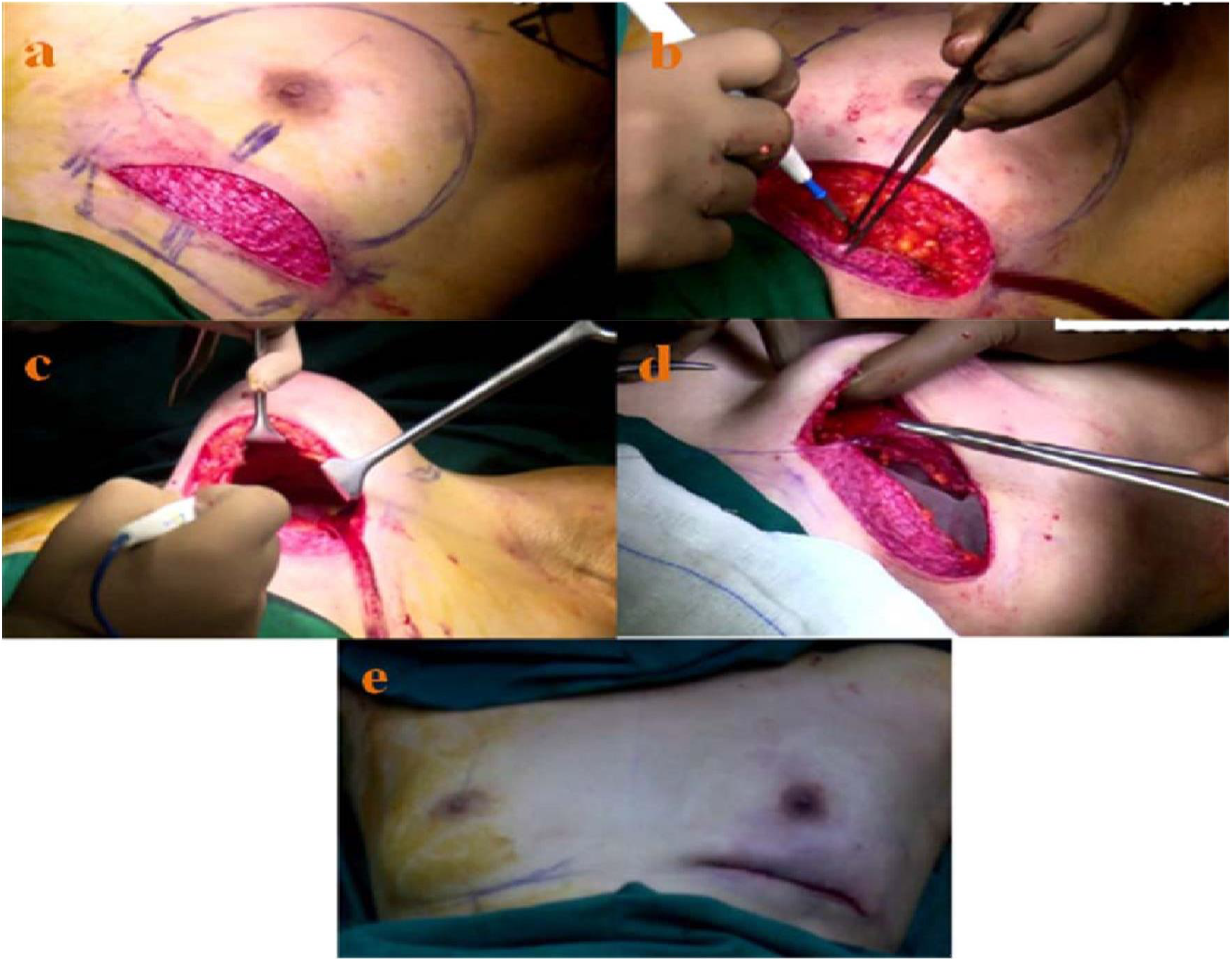
Representative Case Study for A-ALDS based IBRS. 1A. Intraoperative Images a) De-epithelialization of lower dermal flap from IMF to 3 cm onto the breast b) Advancement of lower dermal flap to create of new-IMF by mobilizing skin from lower thoracic and upper abdominal wall c) Formation of subpectoral pocket d) Insertion of implant under pectoralis major muscle e) Completion of IBRS with A-ALDS procedure after skin suturing.

### Study Assessments

Surgical outcomes were assessed by a team of onco-surgeons for post-surgery outcomes. Early complications such as hematoma, seroma, wound dehiscence and wound infection were recorded. Complications were classified as ‘major’ when they required surgical intervention and ‘minor’ when they were managed conservatively. Major immediate complications include implant loss and skin dehiscence that required re-suturing. Minor complications include minor skin/wound dehiscence, minor flap necrosis healing with secondary intention and epidermolysis.

The late complications such as capsular contracture and late infections were also noted. The capsular contracture was assessed using the modified Baker classification system [6]. While, Baker 3 and 4 observations were considered as major complications, Baker 2 observations were considered as minor. We also noted the time between completion of the surgery and start of the adjuvant therapy to ascertain any delays in the adjuvant therapy.

The Patient reported Outcome Measures (PROMs) were used to evaluate patient satisfaction and quality of life after IBRS. To assess PROMs, a standardized Breast-Q questionnaire was utilized. The Breast-Q reconstruction module was divided into multiple independent scales. Higher scores indicate greater patient satisfaction and functionality [7, 8]. PROMs patient interview was conducted by a well-trained interviewer after obtaining informed consent.

Aesthetic outcomes were measured with 5 different variables that included breast reconstruction volume, contour, implant placement, scarring, and appearance of inframammary fold [9]. Post-operative cosmetic assessment was performed by the clinical team during visual inspection of the patient in the sitting position. Photographic data were scored by 3 independent clinical observers using the 3-point scale (Online Resource 4). Two of them were not directly involved with direct patient care. Aesthetic scores of 5 different variables were pooled and analyzed for statistical significance.

### Statistical Analysis

Statistical analysis was performed using Student’s *t*-test and a p-value of <0.05 was considered significant.

## RESULTS

### Representative Case Study

A patient between 20-40 years age group with B-cup breasts and no ptosis presented with a lump in the left breast. On radiological investigations, it showed multicentric tumors in the upper outer quadrant and diffuse microcalcifications. She underwent nipple-areola-sparing mastectomy followed by IBRS with A-ALDS technique. Post-surgery histo-pathological examination revealed that the sentinel lymph nodes were free of tumor (0/3) and with clear tumor margins. The intraoperative, pre- and post-surgery images for this patient are depicted in Figure 1A, 1B, 1C and 1D respectively.

**1B:**
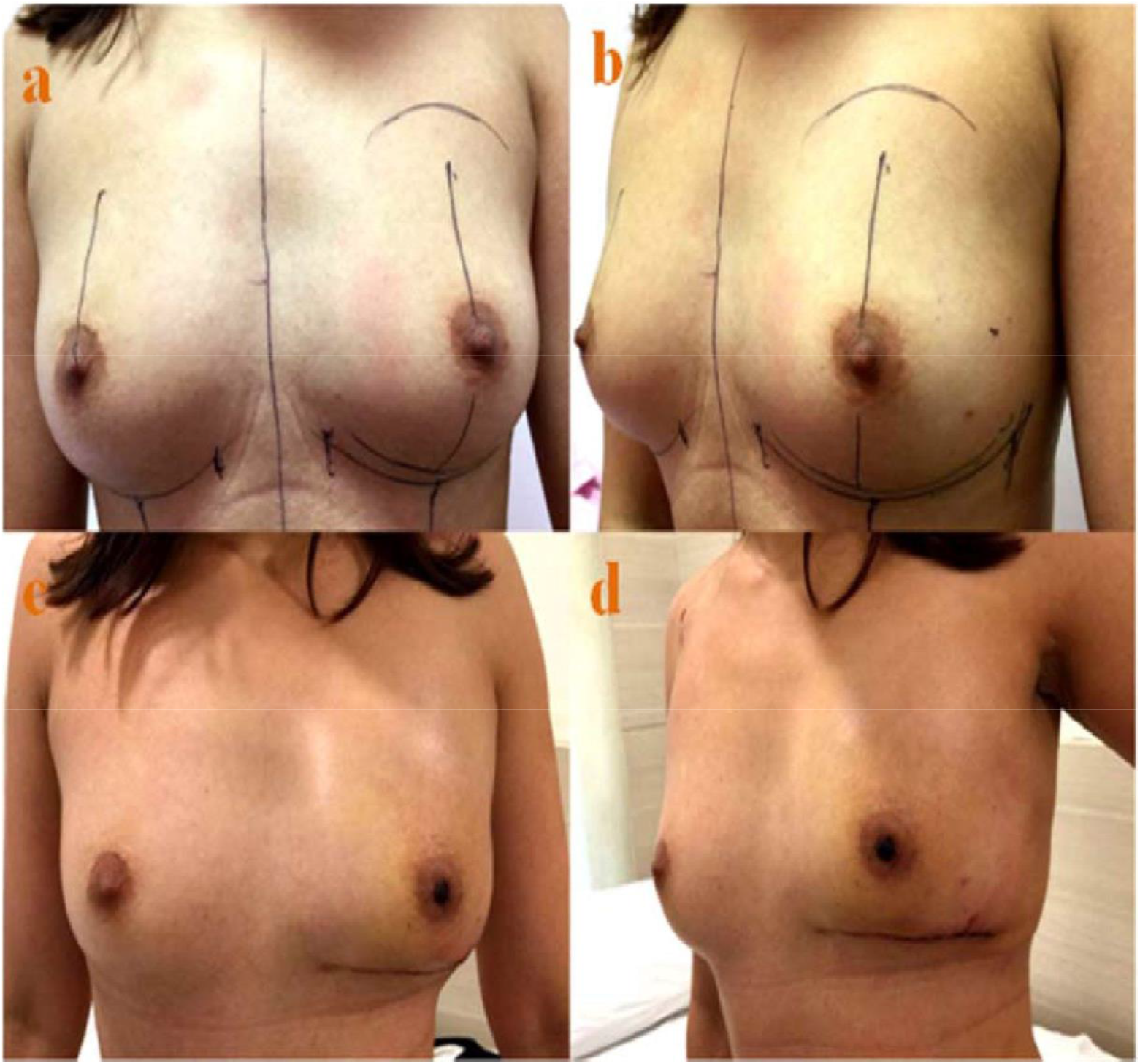
Pre-operative and Post-operative Images. a, b) Pre-operative images c, d) Post-operative images after 1 months of the surgery. Lower pole flattening seen in early phase.

**1C:**
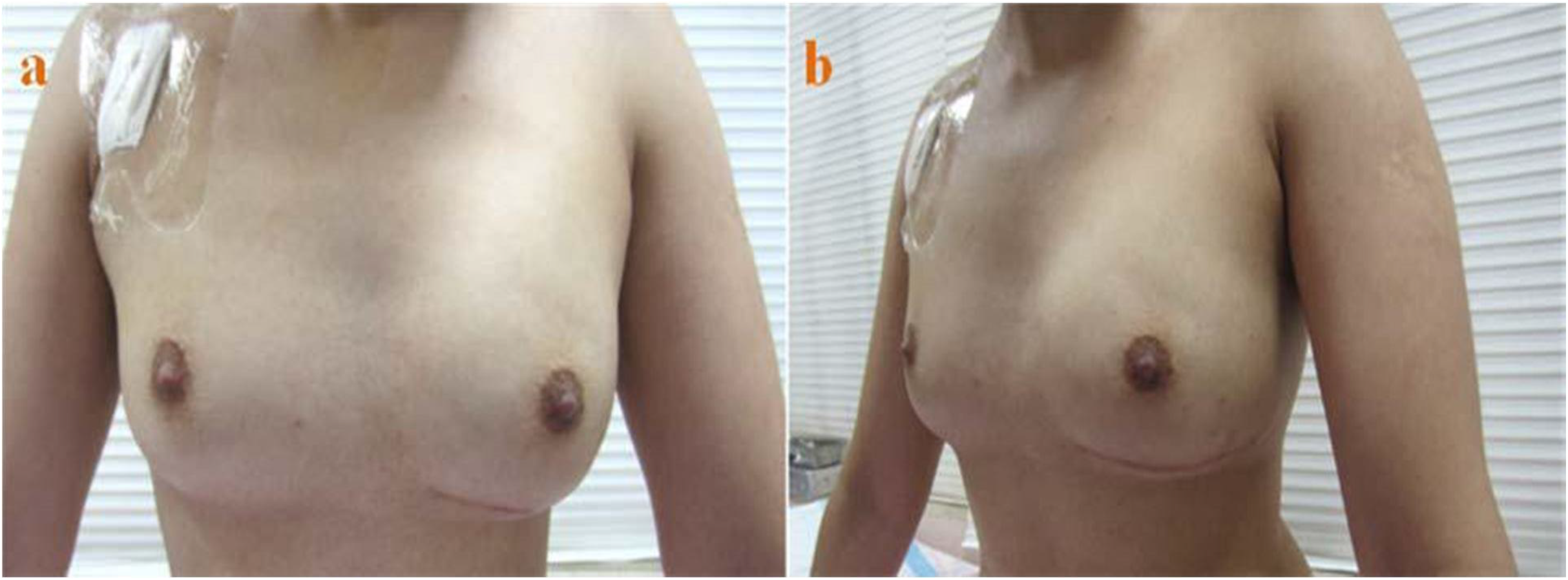
Post-operative Image of the same patient after 3 months follow-up. a, b) Post-operative images after 3 months of the surgery and Lower pole fullness achieved.

**1D:**
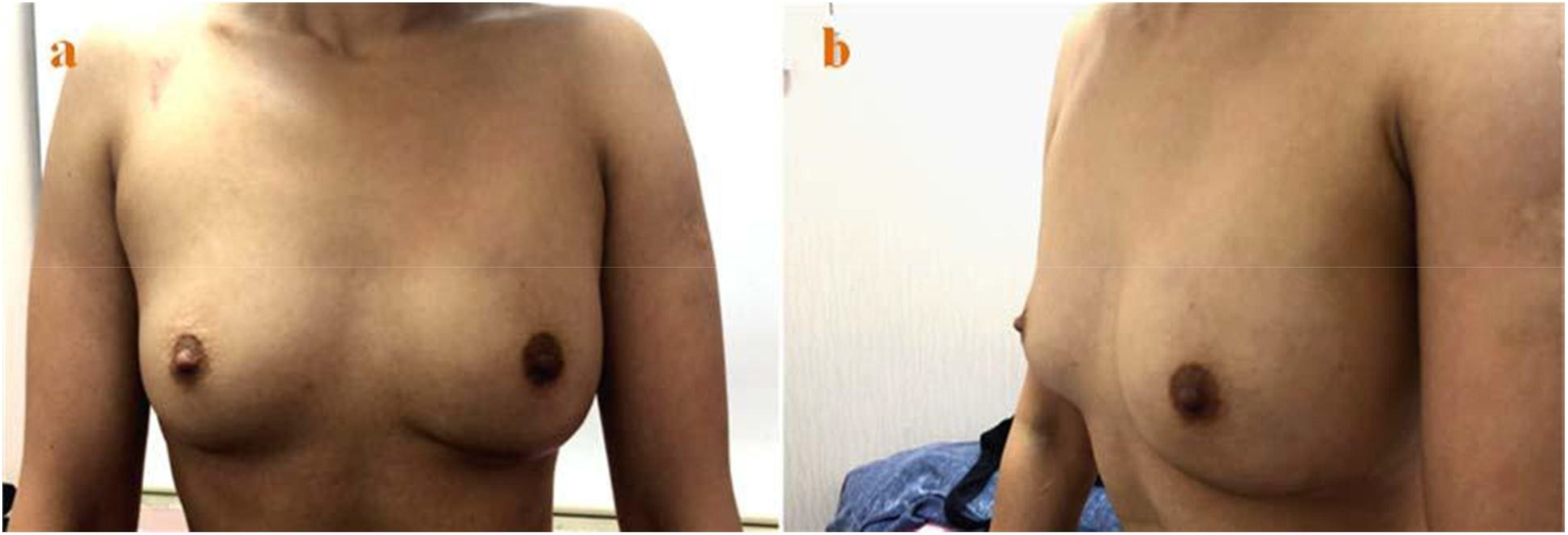
Post-operative Image of the same patient after 1 year follow-up. a, b) Post-operative images after 1 year of the surgery.

### Study Cohort Demography

40 patients with small breasts who completed 12 months post-surgery follow-up were included in the study. Of these, 22 patients underwent implant-based conventional IBRS and 18 patients underwent surgeries with the A-ALDS technique. Out of 40, data of 39 patients was available. Demographic distribution of patients and clinico-pathological characteristics are summarized in Figure 2 and Table 1. None of the patients in our study cohort experienced any delays in their adjuvant therapies.

**Table 1:** Demography and Clinico-Pathological Profile of Study Participants.

| **Variables** | **Data Sources** | **Class intervals** | **Conventional IBRS**<br>**n = 22\***<br>**Number, (%)** | **IBRS with A-ALDS**<br>**n = 18**<br>**Number, (%)** |
| --- | --- | --- | --- | --- |
| **Age (Years)** | Scheduled Interview (Post informed consent) | **<=35** | 4 (18.1 ) | 3 (16.7) |
|  |  | **36-50** | 7 (31.8) | 9 (44.4) |
|  |  | **>=51** | 10 (45.4) | 6 (33.3) |
|  |  | **Mean Age** | 45.6 | 48.2 |
| **Tumor Pathology** | Clinico-Pathological Report (Histo-pathology) | **IDC** | 16 (72.7) | 9 (50.0 ) |
|  |  | **DCIS** | 2 (9.1) | 2 (11.1) |
|  |  | **IDC + DCIS** | 2 (9.1) | 7 (38.8 ) |
|  |  | **Others** | 1 (4.5) | - |
| **ER** | Clinico-Pathological Report (IHC) | **Positive** | 18 (81.8) | 15 (83.3) |
| **PR** | Clinico-Pathological Report (Histo-pathology) | **Positive** | 6 (27.3) | 13 (72.2) |
| **Her2** | Clinico-Pathological Report (Histo-pathology) | **Positive** | 3 (13.6) | 2 (11.1) |
| **TNBC** | Clinico-Pathological Report (Histo-pathology) | ----- | 2 (9.1) | 2 (11.1) |
| **Grade** | Clinico-Pathological Report (Histo-pathology) | **I** | 2 (9.1) | - |
|  | Clinico-Pathological Report (Histo-pathology) | **II** | 15 (68.2) | 15 (83.3) |
|  | Clinico-Pathological Report (Histo-pathology) | **III** | 4 (18.2) | 3 (16.7) |
| **Stage** | Pathology Lab Report (according to AJCC staging guidelines 7<sup>th</sup> ed) | **I** | 9 (40.9 ) | 4 (22.2 ) |
|  | Pathology Lab Report and according to AJCC staging guidelines 7<sup>th</sup> ed | **II** | 4 (18.2 ) | 10 (55.5) |
|  | Pathology Lab Report and according to AJCC staging guidelines 7<sup>th</sup> ed | **III** | 8 (36.4) | 4 (22.2) |
| **Neo – Adjuvant Therapy** | Medical Oncologist consultation and records |  | 4 (18.2) | 4 (22.2) |
| **Adjuvant Therapy** | Medical Oncologist consultation and records |  | 21 (95.5) | 15 (83.3) |
| **Radiation Therapy** | Radiation Oncologist consultation and records |  | 12 (54.5 ) | 8 (44.4) |

\* Data available for 21 patients only.

**Figure 2:**
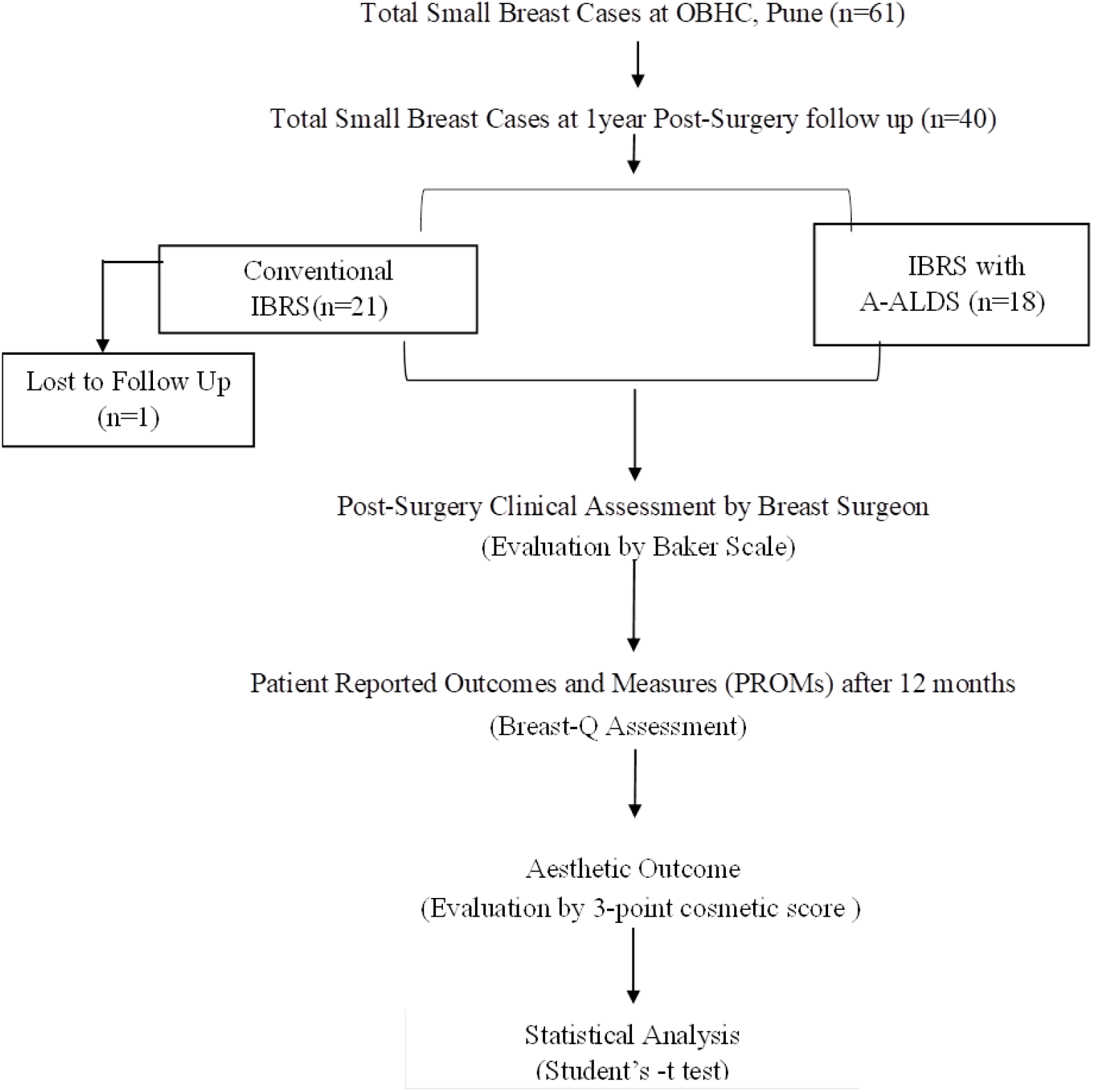
Study Flow Chart : Distribution of Study Participants.

### PROMs and Aesthetic Score

PROMs data was collected from the study participants at 12 months post-surgery with the Breast-Q questionnaire. Out of 40 study participants, 36 (90%) responded to the questionnaire. PROMs data indicated that all study participants irrespective of the type of reconstruction, reported good-to-excellent satisfaction for the breast cosmetic outcomes and psychosocial well-being.

However, we found that the Breast-Q parameters scored higher for A-ALDS patients as compared to conventional IBRS.

The sexual well-being scores were significantly higher for the A-ALDS patients (62.8 ± 21.9 versus 52.2 ± 28.4) in comparison to conventional IBRS procedure (p = 0.0139). In addition, A-ALDS group scored significantly higher for aesthetic score over conventional IBRS group (i.e., 6.7 ± 1.3 versus 4.6 ± 2.3; p=0.0009) (Table 2).

**Table 2:**
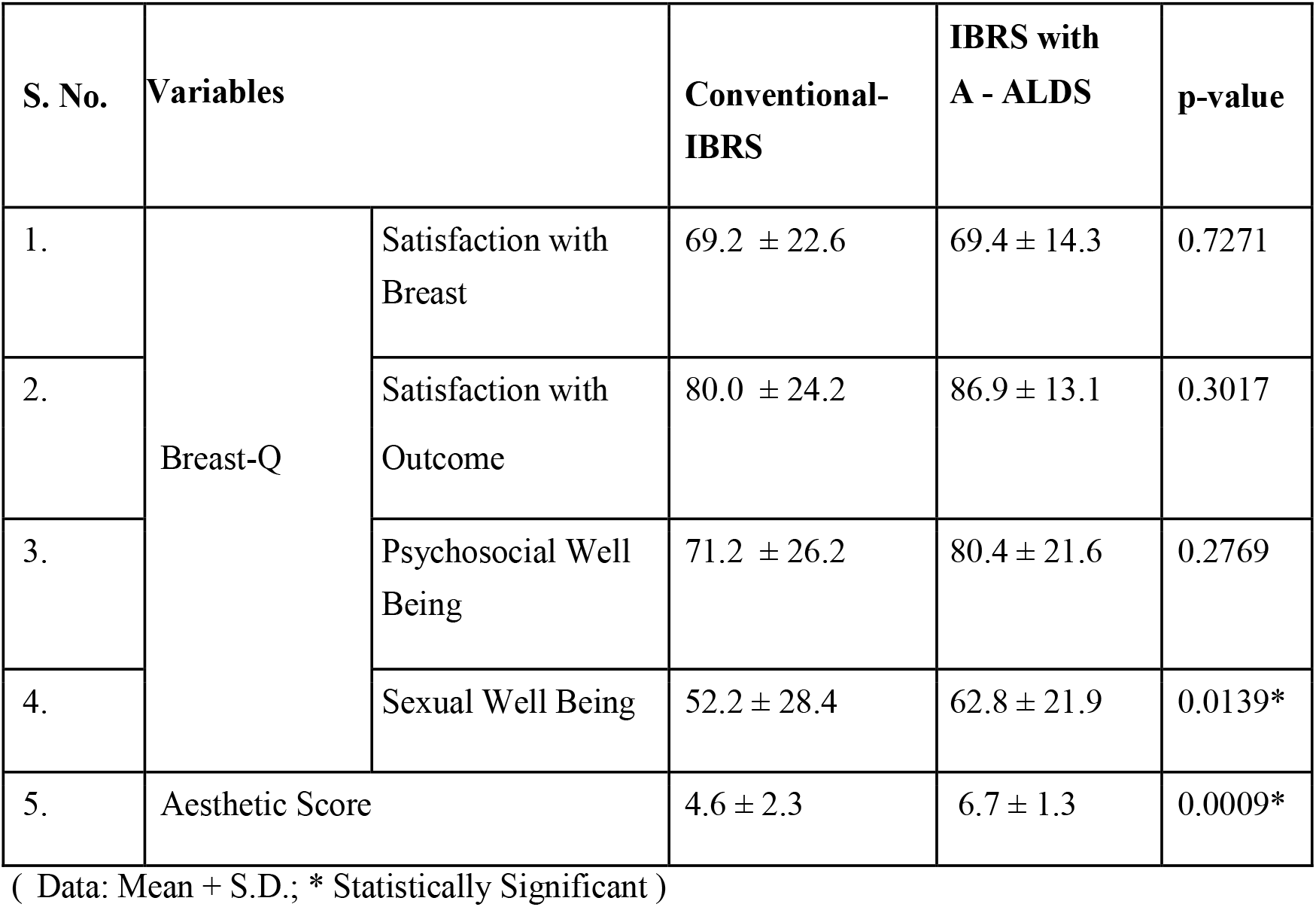
Inter-Group Comparison of PROMs (Conventional-IBRS versus A-ALDS) and Aesthetic Score.

| S. No. | Variables |  | Conventional-IBRS | IBRS with A - ALDS | p-value |
| --- | --- | --- | --- | --- | --- |
| 1. | Breast-Q | Satisfaction with Breast | 69.2 ± 22.6 | 69.4 ± 14.3 | 0.7271 |
| 2. |  | Satisfaction with Outcome | 80.0 ± 24.2 | 86.9 ± 13.1 | 0.3017 |
| 3. |  | Psychosocial Well Being | 71.2 ± 26.2 | 80.4 ± 21.6 | 0.2769 |
| 4. |  | Sexual Well Being | 52.2 ± 28.4 | 62.8 ± 21.9 | 0.0139* |
| 5. | Aesthetic Score |  | 4.6 ± 2.3 | 6.7 ± 1.3 | 0.0009* |

( Data: Mean + S.D.; \* Statistically Significant )

### Assessment of Post-IBRS Complications

No major complications (immediate or /delayed) were observed in the patients that underwent A-ALDS (n=18) procedure. However, out of the 22 patients that underwent conventional IBRS, immediate major complications were observed in 3 patients (13.6 %) with implant loss and delayed major complications in 1 patient (4.5 %) with Grade III capsular contracture (Table 3). No delay in time between completion of the surgery and start of the adjuvant therapy were observed in patients from both groups.

**Table 3:**
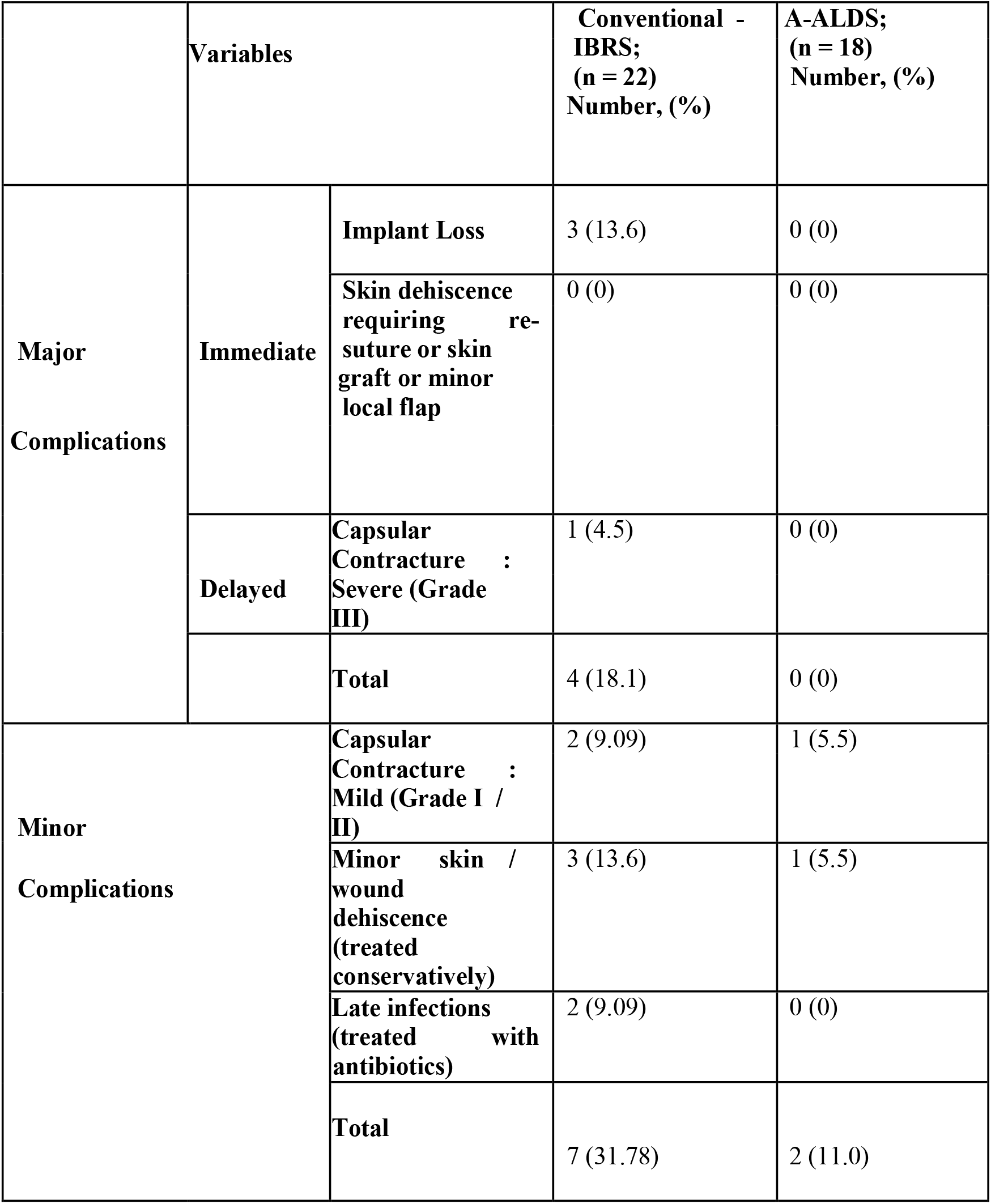
Summary of Post-Operative Complications.

As described above, 18 patients with small breasts from our study cohort have successfully undergone the reconstruction with the novel A-ALDS technique without any major complications. These observations indicate that the use of A-ALDS procedure during IBRS is a safe and feasible technique.

## DISCUSSION

Breast reconstruction in patients with small-breasts is challenging because of insufficient tissue access in the lower pole to create a dermal sling. In this study, we have described in detail a novel A-ALDS technique, which represents an innovative modification to the routine lower dermal sling procedure for application in the reconstruction of small, non-ptotic breasts. In our study cohort, we have observed an early trend towards lower rates of capsular contracture (Grade II and III) and implant loss in A-ALDS patients in comparison to conventional IBRS patients.

A-ALDS provides a stable, double-layered vascularized tissue cover for the implant. This likely allows effective in situ placement of the implant in the breast pocket with robust mechanical stability and helps in symmetrization with the contralateral breast.

The A-ALDS was performed by advancing the de-epithelized dermal sling over the implant by mobilizing the skin and subcutaneous tissue from the lower thoracic and upper abdominal wall. This modification results in the recreation of a well-defined IMF to facilitate symmetry *vis-svis* shape and size with contralateral breast. In our technique, the A-ALDS flap was sutured with pectoralis major/serratus anterior muscle to provide desired expansion to the lower pole for implant placement that provides mechanical stability to the implant. The A-ALDS flap allows the expansion of the inferior pole of the pocket and prevents high riding of the implant. Hence, this modification provides a good contour and natural shape and symmetry to the breast. It is well reported that thoraco-epigastric flaps maintain vascularity from perforators of the intercostal, lumbar, epigastric arcade and inferior epigastric arteries. As a result, these flaps have been previously used in correcting mastectomy defects by small volume replacements [10].

We now present evidence of successful use of A-ALDS flap in breast reconstruction without detaching and advancing it by preserving the vascularity. In our cohort, A-ALDS patients demonstrated significantly higher aesthetic scores (6.7 ± 1.3 versus 4.6 ± 2.3, p=0.009) compared with conventional IBRS after a 12 month follow-up.

In addition to optimal post-surgery outcomes, patient acceptance of the A-ALDS technique is equally important. The PROMs (Breast-Q) data from our study indicates significantly higher satisfaction with sexual well-being in patients who have undergone reconstruction with the A-ALDS technique. Other parameters such as satisfaction with breast, satisfaction with outcome and psycho-social well-being showed a positive trend in favor of A-ALDS-based reconstruction.

Based on these observations, we hypothesize that use of A-ALDS flap with an implant may provide a viable, vascularized tissue cover to the implant. This flap lowers the risk of implant exposure, thereby, lowering the rates of implant loss and capsular contracture. This well vascularized autologous flap is expected to render the tissue environment more favorable for implant placement and improve wound healing, thereby reducing the risks of fibrosis, capsular contracture, wound breakdown, infection and may reduce the implant related complication after RT [11]. By extension, the higher complication rate in our study patients who have undergone conventional IBRS may be attributed to the thin fascia covering the implant, which may predispose the implant to exposure in case of infection or necrosis.

In low-resource settings such as India, ADMs are not yet available. This prompted us to innovate the breast reconstruction technique for women with small breasts using the A-ALDS technique. Indeed, our collective results indicating superior cosmetic scores, lower rates of immediate complications and trend towards better patient acceptance after A-ALDS based reconstruction are encouraging. It is conceivable that our novel A-ALDS technique may provide a cost-effective alternative to ADM based IBRS. The economic advantage of this ALDS technique is apparent as it can be performed in a single setting without compromising the patient outcomes and obviating the need for contralateral procedure.

Our main aim was to report the surgical details of the innovative A-ALDS based IBRS procedure in small or minimally ptotic breasts. We propose that the A-ALDS based IBRS procedure may be routinely employed in the patients with small breasts that will ensure a good cosmetic outcome with minimal early complications, no implant loss and lesser capsular contracture rate with an overall positive impact on quality of life.

Despite the important findings, our study has few limitations. This study represents the post IBRS follow-up data only for a period of 12 months. To substantiate the promising clinical and PROMs observations, long-term follow-up i.e., 3 to 5 years post-surgery in the same cohort is needed. Secondly, this study represents a single-institutional cohort in which all A-ALDS procedures were performed by the same surgical team which may represent investigator bias. To validate our observations and minimize bias, this study needs to be replicated in multi-centric settings.

## CONCLUSION

In conclusion, our study reports application of a novel A-ALDS technique for implant-based breast reconstruction in small, non/minimally ptotic small breasts. Preliminary observations from post-surgery evaluations and PROMs demonstrate that A-ALDS technique may potentially reduce post-surgery complications, improve aesthetic outcomes and improve overall patient satisfaction. This technique may serve as a cost-effective alternative to ADM-based reconstruction in low resource settings. Long-term follow-up and study replication in other breast surgery units will be necessary for further substantiating our early observations.

## Data Availability

The study investigators would like to state that all data related to the manuscript will be available for further review on request after consultation with institutional ethics committees

## Abbreviations (According to Appearance in the Text)

ALDS: Autologous Lower Dermal Sling
IBRS: Immediate Breast Reconstruction Surgery
BC: Breast Cancer
IMF: Infra-mammary Fold
ADMs: Acellular Dermal Matrices
A-ALDS: Advanced-Autologous Lower Dermal Sling
PROMs: Patient Reported Outcome Measures

## DATA AVAILABILITY

All data will be available and can be shared on request. Supplementary tables and figures are provided in support of data presented in the manuscript.

## ACKNOWLEDGMENTS

The study authors would like to thank all participants who consented to participate in this study. We also acknowledge Bajaj Auto Ltd. for providing support to research activities at PCCM (Grant #PCCM529). We express our appreciation to Dr. Madhura Kulkarni, Dr. Dhara Patel and Dr.Sneha Joshi for their help in editing the manuscript. We are grateful to the MAPI Research Trust for permission to use BREAST-Q (http://www.mapitrust.org). We would also like to thank the management and staff of Ruby Hall Clinic, Pune where all surgeries were performed.

## ETHICAL APPROVAL

Study approval was granted by the Independent ethics committee of Prashanti Cancer Care Mission. Independent Ethics Committee of Prashanti Cancer Care Mission (DCGI/CDSCO Registration Number : ECR/298/Indt/MH/2018 (dated May 14 2018)

## CONSENT

All subjects gave their consent for the use of their personal and medical information including images in the publication of this study.

## FOOTNOTE

A part of this work was presented as a poster at the Breast Cancer Coordinated Care conference in Washington, DC on March 1-3, 2018.

## SUPPLEMENTARY MATERIAL

### Supplementary Videos

**Koppiker etal - A-ALDS Video 1**.**mp4 : Pre-Operative Marking and Creation of A-ALDS Flap**

Pre-Operative markings and skin de-epithelisation to form A-ALDS flap.

**Koppiker etal - A-ALDS Video 2**.**mp4 : Mastectomy**

Mastectomy is carried out with the removal of breast tissue.

**Koppiker etal - A-ALDS Video 3**.**mp4 : Correction of Completely Vascularized Pocket with A-ALDS Technique**

Implant is placed in sub-pectoral pocket and advancement of A-ALDS flap. Pectoralis major and serratus anterior muscle are sutured to de-epithelialised A-ALDS to create a complete vascularized cover.

## SUPPLEMENTARY TABLE

### Aesthetic 3-Point Outcome Score

| **Category** | **0** | **1** | **2** |
| --- | --- | --- | --- |
| **Volume** | Marked discrepancy relative to the contralateral side | Mild differences in Volume relative to the Contralateral Side | Minimal Differences in volume relative to The Contralateral Side |
| **Contour** | Marked contour deformity or shape asymmetry | Mild differences in Contour deformity or Shape Asymmetry | Minimal Differences in contour deformity or Shape Asymmetry |
| **Placement** | Marked displacement of breast mound | Mild Displacement of breast Mound | Minimal Displacement of breast Mound |
| **Scarring** | Hypertrophic scars and evident contracture | Fair scarring (poor color match or wide scars without Hypertrophy Or contracture) | Thin scars |
| **Infra-Mammary Fold** | Poorly defined infra-Mammary fold | Defined but Asymmetrical infra-Mammary Fold | Symmetrical infra-Mammary Folds |
| Nguyen et al. *Eplasty* 2012; 5:486–492. |  |  |  |

